# Quality of maternal care at public health centers of Siltie zone, Central Ethiopia 2022

**DOI:** 10.1101/2024.01.26.24301840

**Authors:** Abdulfeta Shafi Mohammednur, Neima Ridwan Abdu

**Author notes:** corresponding author: Email address (AS). These authors contributed equally to this work.

## Abstract

**Introduction:** Quality of care is “the extent to which health care services provided to individuals and patient populations improve desired health outcomes. In order to achieve this, health care must be safe, effective, timely, efficient, equitable and people-centered (WHO). Quality care is crucial in ensuring that women and newborns receive interventions that may prevent and treat birth-related complications. This paper assessed the quality of maternal care at public health facilities of Siltie zone, Central Ethiopia, from March 1 up to April, 15, 2023.

**Method:** facility based Cross sectional study design was conducted. Under this study totally 437 individuals’, 367 mothers and 70 health care providers in 23 health center were interviewed. All woreda and city administrates were included in the study contributing at least one health center. The data collection tool was adopted from health sector transformation in quality in Ethiopia (HSTQ). Specific health center maternal health care quality score was obtained by the total score of the health center performed divided by the total score expected (excluding NA quality measures) and the result was multiplied by 100%.

**Result:** Overall quality of sanitation facility in health center is 71% and the overall quality of care service based on the health care provider is almost 80.7%. Quality of prenatal health care service is 83%, Labor and Newborn care is 87%, postpartum care 87%and Facility inventory 77%. The overall quality of maternal care is 82%.

**Conclusion:** Quality of maternal and newborn health care service is good. Further efforts are needed to improve availability of necessary equipment’s, electricity and water supply.

## Introduction

Much progress has been made during the past two decades in coverage of births in health facilities; however, reductions in maternal and neonatal mortality remain slow. With increasing numbers of births in health facilities, attention has shifted to the quality of care, as poor quality of care contributes to morbidity and mortality (1).

Quality of care means many things to different people and there is no single universally accepted definition(2). The world health organization (WHO) definition of quality of care is “the extent to which health care services provided to individuals and patient populations improve desired health outcomes. In order to achieve this, health care must be safe, effective, timely, efficient, equitable and people-centered”(1).Another definition of quality of care in maternity services involves providing a minimum level of care to all pregnant women and their newborn babies and a higher level of care to those who need it. This should be done while obtaining the best possible medical outcome, and while providing care that satisfies women and their families and their care providers(2).

Quality care is crucial in ensuring that women and newborns receive interventions that may prevent and treat birth-related complications. As facility deliveries increase in developing countries, there are concerns about service quality(3).Measurement of the quality of routine intra-partum and immediate postpartum services is essential in ensuring the delivery of appropriate interventions to reduce maternal and newborn mortality and morbidity(3).

The quality of care received by mothers and babies in developing countries is often reported as poor. Yet efforts to address this contributory factor to maternal and newborn mortality have received less attention compared with barriers of access to care. Quality care should thus lie at the core of all strategies for accelerating progress towards MDG 4 &5 (4).

The study on assessment of the quality of maternal care in health facility is very scares in Ethiopia. Effective prevention and management of conditions in late pregnancy, childbirth and the early newborn period are likely to reduce the numbers of maternal deaths, ante partum and intra-partum-related stillbirths and early neonatal deaths significantly. Therefore, improvement of the quality of preventive and curative care during this critical period could have the greatest impact on maternal, fetal and newborn survival (1).

Despite the fact that quality maternal care is essential for further improvement of maternal and child health, to the best of researchers’ knowledge, there is no study done in Siltie zone Central Ethiopia in general. Therefore, this study aims to fills this gap and assesses the quality of maternal care in Siltie zone at health center. Specifically, assess the quality of prenatal service, qualities of intra-partum and newborn care service; qualities of postpartum service and assess availability of resource inventory in Siltie zone at public health facilities.

### Research question

What is the level of quality of maternal care in Siltie zone?

## Methods and material

### Study area and period

The study was conducted in Siltie zone from March 1 up to April, 15, 2023, Central Ethiopia region. This zone had 10 woreda and 5 city administrators with 35 health centers and four hospitals. The study included all woreda and city administration. Totally 23 health center were randomly selected for study with Minimum of two health centers per each woreda. Three health centers were considered in alicho wirirro and silti woreda. But, Werabe and Alkeso health center included from Werabe city administrator.

### Study Design

Facility based cross sectional study design was conducted.

### Study population

All pregnant women who had both ANC visit and admitted for delivery at selected public health facilities of Siltie zone were used as source of population for this study. Sampled pregnant women undertake ANC visit and admitted for delivery at selected public health facilities during the study period were study unit

### Inclusion and exclusion criteria’s

All pregnant women who had ANC visit before in the health center and admitted to give birth during data collection period and can give consent included. All pregnant women having complication like postpartum hemorrhage, pre-eclampsia or eclampsia were excluded. Women admitted at an advanced stage of labor (third stage) with frequent (≤5 min apart) and intense uterine contraction that would prevent them from providing valid informed consent were excluded from the study. Mothers who are very sick and referred to another health facility also excluded.

### Study variable

Quality of maternal health care service in Siltie zone

### Sample Size

A single population proportion formula used to estimate sample size of mothers to be investigated, with assumptions of 5% margin of error, 95% confidence, and a non-response rate of 10% and the proportion (p) of quality of maternal health care service.

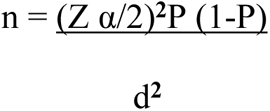

Where n= total required sample size, Z α/2= critical value 1.96 at 95% confidence interval P= 0.5, proportion of quality of maternal health care service and q=1-0.5=0.50 d = 5%

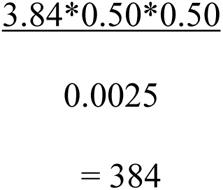

Proportion of Quality of maternal care service (p=0.5) with 10% of non-response rate the final Sample size was 423.

### Data Collection tools, Procedure and data quality

Data collection tool was adopted from Ethiopian health sector transformation in quality (HSTQ) guide line and maternal and child birth integrated program (MCBIP).The reliability of the tool assessed by Cronbach’s alpha which equals to 0.92 is. The tool was adopted to assess the quality at prenatal, intra-partum and new born, postpartum and facility inventory. Totally, 100 quality measures item from six quality standards were adopted to assess quality of maternal care received from national and WHO maternal and new born health care transformation. Four types of data collection methods were used to collect data from health centers. (1) Interview with MCH department, health care providers and clients (2) client chart review, (3) resource facility inventory which recorded the availability of essential drug, supplies, equipment, guidelines and infrastructure (5) Structured clinical observation checklist for labor and delivery (L&D) and newborn care. The resources inventory and protocols conducted once per heath center and in addition to observation include interviews with MCH department. The facility inventory checklist focused on collecting information on infrastructure conditions and verifying the availability of medications, supplies, equipment and protocols

A pretest was conducted on 22 women and 4 health care providers on Butajera health center and necessary modifications made on the tool based on the findings. One-day training was provided for data collectors and supervisors about the purpose of the study and techniques of data collection.

A total of 23 midwives and/or nurses as data collectors and 5 supervisors recruited for data collection. To retain quality of the data, supervisors supervise data collectors and the collected data timely. Exit interview conducted among postnatal women using the questionnaire.

### Sampling Technique

Stratified sampling technique was used to select women from health centers. Those are West Azernet, East Azernet woreda, Alicho wirirro woreda, Hulbarage woreda, Mito woreda, Sankura woreda, dalocha woreda, silti woreda, East silti, Lera woreda, Lanfuro woredas and Werabe and Alkeso health center from Werabe city were considered.

Total sample of 423was allocated to each health Center according to average number of monthly delivery report per month in health facility. Consecutively, all women qualified to study under postpartum ward randomly selected and interviewed before leaving health center until desired sample size obtained.

### Data Processing and Analysis

Data was collected by using kobo toolbox and exported to SPSS version 21 for analysis then explored for missed values, outliers, and any inconsistencies. Microsoft excel was used to analyze the data. Descriptive statistics, text narration, and tables used to present the results. Descriptive statistics like percentage, graphs and descriptive summaries were used to describe the study variables. Specific health center maternal health care quality score obtained by the total score of the health center performed divided by the total score expected (excluding NA quality measures) and the result multiplied by 100%.

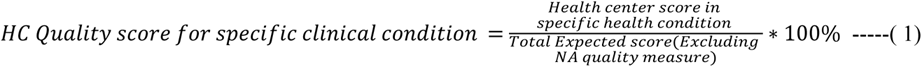

Unless specific direction is provided for a specific quality measure, the following general guidance was used for all quality measures requiring client interview, staff interview and chart review

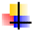 For those quality measures requiring client interview for verification, select 5-25 clients per health center leaving the facility after service use during data collection period.
  ➢ Conduct exit interview for the required information.
  ➢ Score each client response from 2 if the criteria met
  ➢ Score 0 for each client response if the criteria is unmet
  ➢ NA for each specific case not identified
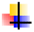 For those quality measures requiring staff interview for verification, interview 2-5 midwife staffs (as specified in the remark section) working in the facility
  ○ Conduct interview/ skill demonstration for the required information.
  ○ Score each staff response from 2 if the criteria met
  ○ Score 0 for each staff response if the criteria is unmet
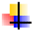 **Trace the charts from the medical record room**
  ▪ Verify if the required information is documented in the chart
  ▪ Each chart scored from 1 or 0 depending on the presence or absence of the information respectively, and totally the quality measure scored.
  ▪ If the documented information is not legible, that specific chart given a score of 0
  ▪ Absence of documentation is taken as the service was not provided
  ▪ NA for each chart for which the specific clinical condition is not identified.

## Result

### Socio-demographic characteristics of client and health care provider

Among 70 health care provider 46 (66%) were female and 24 (34%) were male. Most of health care providers 39 (56%) had diploma and 31 (44%) were degree holder. Among health care providers only 8 of them have one-year experience and 59 (88%) had more than or equal to two years’ work experience.

The mean age of health care provider was 28.3 year with min 20 and max 45. Average age in year for women is 28.98 (SD=5.74). Most of women and health care provider were in 20-30 age categories which account about 30.6% and 48.6% respectively. Most of women were from rural area 287 (78.2) and more than 47% of mother were home-maker. Related to maternal education about 42.2% of mother had primary education and only 6% of them had more than secondary education.

### Facility inventory

The facility inventory for health care service was measured by using 38 questions adapted from national HSTQ. The question relates with number of bed, essential equipment availability, essential drug availability, electric supply, lab test, sanitation service, waste management and availability of water in sufficient quantity.

Two out of twenty-three health centers had no functioning first stage bed. The rest 91% had at least two first stage beds and minimum of one second stage bed at labor.

Related to medicines and equipment supplies, all labor ward had an emergency drug cabinet that has labeled essential drugs and stock management in place and a clear communication channels is present to reach staff on duty at all times in all health center.

All the necessary equipment’s needed for newborn resuscitation were available only in 4 (13%) health centers. Relevant guide lines needed in the labor and delivery room are available in 21 (91%) health centers. A dedicated area was present in labor and delivery area for resuscitation of newborns (Newborn Corner).

Only 4 (17.4%) health centers had all Essential elements (Essential drug, equipment’s and lab test) in the labor ward in sufficient quantity during data collection period. 30.4% of health center had all essential drugs and equipment’s. however, one drug missed in 26.1%, one essential equipment missed in 17.4% and one lab test missed in 60.9% of health center. All Essential drugs, Essential equipment’s and Essential lab tests are available in the labor ward at all times in sufficient quantity in 30.4%, 30.4% and 26.1% respectively. But, at least two Essential drugs, two Essential equipment’s and two Essential lab tests are missed in health center in the labor ward about 43.5%, 52.2% and 13.0% respectively.

Only, 54% of Neonate were given vitamin K1mg, TTC eye ointment and vaccinated with BCG and OPV0 and basic Neonatal care is given as per national recommendation 91% of newborn.

### Sanitation and hygiene facility

All Health center sanitation elements are satisfied in only 13% of health center and one element missed in 48% of health centers. Overall quality of sanitation facility in health center is 71% **Table 2** below.

**Table 1:**
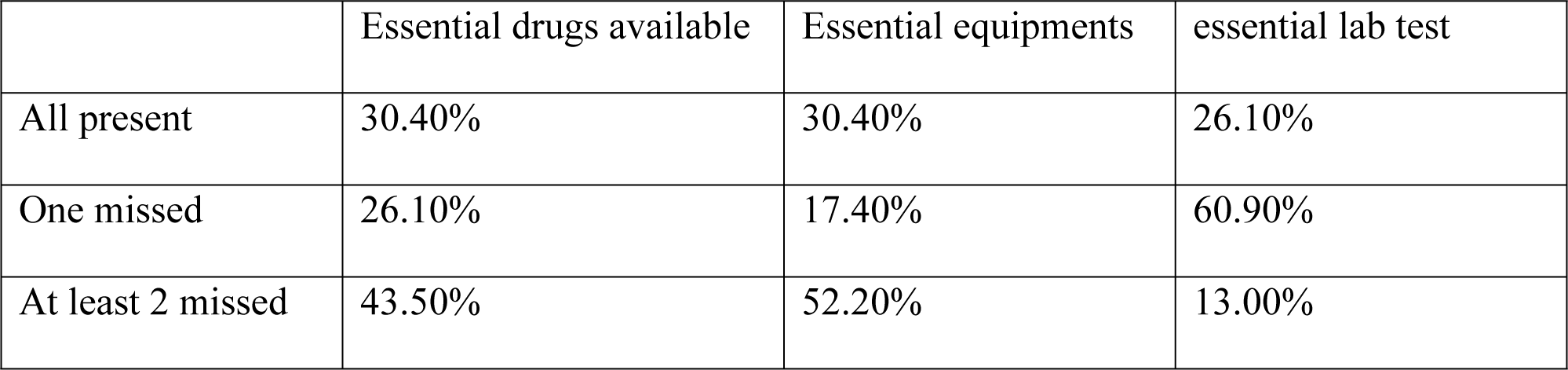
Availability of essential drug, equipment and lab tests.

|  | Essential drugs available | Essential equipments | essential lab test |
| --- | --- | --- | --- |
| All present | 30.40% | 30.40% | 26.10% |
| One missed | 26.10% | 17.40% | 60.90% |
| At least 2 missed | 43.50% | 52.20% | 13.00% |

**Table 2:** Availability of sanitation and hygiene facility elements (N=23 health center)

| SNO | Sanitation facilities elements | Percentage | Over all Quality |
| --- | --- | --- | --- |
| 1. | Appropriately illuminated at night | 83% | 71% |
| 2. | Accessible to people with limited mobility | 83% |  |
| 3. | Gender separated for staff and attendants | 87% |  |
| 4. | Hand washing stations with soap and water | 57% |  |
| 5. | sanitation facilities are/Adequate number (at least 1 latrine per 20 users for inpatient settings) | 87% |  |

### Health care provider as measure of quality

70 health care providers (midwifes) from 23 health centers were interviewed during study period. 12question related to health care provider were adapted from HSTQ. The overall quality of care service based on the health care provider is almost 80.7%. Nearly, 77.9% of HCP demonstrate clearing their hands correctly as per the WHO5 and 66% of care providers highly satisfied with working environment and support of health center management **Table 3**.

**Table 3:** Quality of health care service based on the health care providers.

| SN | Quality elements | Percentage |
| --- | --- | --- |
| 1. | Satisfaction with water, sanitation and energy service | 52.9% |
| 2. | HCP Demonstrate cleaning their hands correctly as per the WHO 5 | 77.9% |
| 3. | Availability of PPE in need and the health center management is supportive of all inquiries | 77.9% |
| 4. | Highly satisfied” with your job in relation to the working environment and support of health center management | 66% |
| 5. | HC provider prepare 0.5% Chlorine solution | 66.2% |
| 6. | Now how to process used instruments (instrumental processing) | 100 |
| 7. | Demonstrate skills of basic neonatal resuscitation | 94.1% |
| 8. | Staff can describe PPH management Adequately | 100.0% |
| 9. | Describe Eclampsia management adequately | 98.5% |
| 10. | Bi annual appraisal of all staff and a mechanism of recognizing high performing workers is in place | 97.1% |
| 11. | An enabling supportive environment for professional staff development is in place through. | 89.7% |

**Table 4:** Quality of maternal and newborn care service based on pregnancy period and facility.

| SNO | Category | Quality score |
| --- | --- | --- |
| 1. | ANC/prenatal health care service | 83% |
| 2. | Labor and Newborn care | 87% |
| 3. | Postpartum care | 87% |
| 4. | Facility inventory | 77% |

### Quality of maternal health care service in Siltie zone health sector

Under this study to measure maternal health care service in Siltie zone 11 question for ANC/Prenatal care, 31 question for Labor, 17 question for postpartum, 9 question for newborn and 38 question for facility inventory was adapted from health sector transformation in quality in Ethiopia. The quality of health care service for antenatal/prenatal maternal health care service is 83% while 87% for labor and newborn care. Likewise, quality of Health care service for postpartum and Facility inventory is 88% and 77% respectively.

### Overall quality of maternal health care service in Siltie zone health sector

Based on 367 mother and 70 health care provider, Quality of maternal health care service in Siltie zone based on six standards or service given at antenatal care, intra-partum/ labor, postpartum and newborn care service based on adapted hundred questions is 82%.

## Discussion

This study assessed quality of maternal health (prenatal, intra-partum and newborn care, post-partum and facility inventory) care service in Siltie zone health centers. The study result shows quality of maternal health care service is good.

Quality of antenatal care service in Siltie zone is 82% which is much higher than results obtained in Hosanna town in Hadiya zone (31.18%)(6), Sidama region (41.2%)(7), Bangladesh 55% (8)and Bahirdar special zone 52.3% (9). The difference between previous study and present study may be due difference in tools used to assess the quality service, collaborative work of Siltie zone health sectors with Werabe comprehensive and specialized hospital. Hence, quality is multidimensional and our quality assessment tool was adapted from HSTQ with weight as it is. So that, the weighting itself may be cause for difference in quality. In addition, difference might be the experience of the health care providers and availability of facility inventory. The facility supplies are the most expected quality compromiser in health centers. Availability of appropriate working system and physical environment with adequate working guidelines, utilities, medicines, supplies and equipment for providing quality maternal health services is crucial and it accounts 80% in Siltie zone health centers.

This study result shows that postnatal maternal and newborn health care service in Siltie zone is 87%. But, contrast to study done using Ethiopia Mini Demographic and Health Survey 2019 data finding the percentage of participant who score quality care by using EDHS 2019 score is 46% in Addis Ababa which is the highest and minimum score in Harari region 10.7% (10). But, the quality maternal health care service of the other region is much less than Addis Ababa(11). In addition to this, our findings are greater than study done in Bangladesh (8). This may be Siltie zonal health office striving for ending preventable maternal mortality (EPMM) by 2030, every country should reduce its maternal mortality ratio (MMR) by at least two thirds from the 2010 baseline, and no country should have an MMR higher than 140 deaths per 100 000 live births (twice the global target) (5).

This study revealed that only 54% Neonate is given vitamin K1mg, TTC eye ointment and vaccinated with BCG and OPV0. But study done in four regions revealed that 38% of newborns received vitamin K (12). Available of newborn resuscitation bag and mask device is 90%. This is similar with Study in Nepal including governmental hospital, private, primary health center and health post. Availability of newborn resuscitation bag and mask device is 100% in primary health care and governmental hospital (13) and 80% in Ghana (14). Nearly 99% newborns were breastfed within 1hour afterbirth, 54% Neonate is given vitamin K1mg, TTC eye ointment and vaccinated with BCG and OPV0 and 94% newborns get their umbilical cord clamped after 1–3 min of birth. 98% Initiate breast feeding within 1 hour after delivery and 63 % health TTC eye ointment (14).

All Essential drugs, Essential equipment’s and Essential lab tests are available in the labor ward at the time of study period in sufficient quantity was 30.4%, 30.4% and 26.1% respectively. Study done in four regions revealed that essential drugs availability score was 74% and only 3 health facilities with all essential drug inputs available(12). The mean infrastructure availability score was 83.6% which is similar with our study which is 77%. input is 62%, (12) process 48% and output 48% but this study shows that quality ranges from 77% to 87%.

## Conclusion and recommendation

➢ Quality of maternal health care is relatively good for prenatal, labor, postnatal and newborn care services. However, Health care provider less satisfied with Supply of facility inventory electricity, water and backup resource limited and sanitation elements.
➢ Availability of necessary equipment’s needed for newborn resuscitation and PPE is low.
➢ Some health facilities missed screens or curtains b/n each beds to ensure privacy and complaint handling office for handling compliant of mothers and their families.
➢ Equipment’s and drugs needed for newborn resuscitation and newborn care is low.
➢ Quality of Sanitation and hygiene is very low. So that the zonal and regional health bureau and NGOs should have to work to increase sanitation facility and supply.
➢ In case of power cut, generator or alternative energy source is not automatic or cannot start with in short period. Hence, concerned body should solve this problem.
➢ Stakeholders should have to work on all health centers to have adequate backup water source when there is interruption from the main source

## Data Availability

Data Access / Ethics Committee (contact via the corresponding author address) for researchers who meet the criteria for access to confidential data

## Abbreviation

NGO: none governmental organization
WHO: world health organization
ANC: Antenatal care
PPH: postnatal care
MCBIP: maternal and child birth integrated program
MMR: maternal mortality rate
TTC: tetracycline
PPE: personal protective equipment
NA: Not available
HSTQ: health center transformation in quality
HCP: health care provider
NA: Not Available

## Acknowledgment

We would like to thank Werabe University for supporting this research work. We wish to express our deep appreciation to all health care providers, Staff, data collectors and women for their time and genuine response to the overall success of this study.

## Author contribution

Both authors and co-author have designed the study, develop data collection tool. Both supervise data collection, analyze, interpret result and draft, and revised critically and finally approved Manuscript for publication.

## Funding

This research was partially supported by Werabe University. However, the University has no any role in this paper other than funding.

## Availability of data and supplementary material

data will be available upon the reasonable request. Reasonable request can be made to corresponding author

## Ethical approval and consent of participate

Ethical clearance was obtained from Werabe University ethical and review board committee and written permission letters from three woreda were granted. Administrators of selected woreda health center and hospital were informed about the objective of the study. Ethical committee approved verbal consent (meeting number WRU/RPD/09/2022 and protocol number: GOV/WRU/CMHS/MIDW/2022/1521) because most of participant in study area were never attend school. All participants were informed about the purpose of the study. They were informed that, their participation is voluntary and can withdrawal at any time without permission and penalty. Verbal consents were obtained by asking women whether they agree to participate in the study.

## Computing interest

Author has no any computing interest

## Notes

### Competing Interest Statement

The authors have declared no competing interest.

### Funding Statement

Yes

### Author Declarations

Werabe University review and ethical board commitee

